# How has the COVID-19 pandemic impacted on smoking and nicotine dependence among people with severe mental ill health? Analysis of linked data from a UK Closing the Gap Cohort

**DOI:** 10.1101/2021.02.10.21251467

**Authors:** Emily Peckham, Victoria Allgar, Suzanne Crosland, Paul Heron, Gordon Johnston, Elizabeth Newbronner, Elena Ratschen, Panagiotis Spanakis, Ruth Wadman, Lauren Walker, Simon Gilbody

## Abstract

Smoking rates are higher for people who use mental health services which contributes substantially to health inequalities. Smoking can lead to worse COVID-19 outcomes, yet it remains unclear whether smoking has changed for people who use mental health services. We examined smoking patterns in a large clinical cohort of people with SMI before and during the pandemic. We found high levels of nicotine dependence and heavier patterns of smoking. Whilst some people had reported quitting, it is likely that smoking inequalities have become further entrenched. Mental health services should seek to mitigate this modifiable risk and source of poor health.

## Background

People with severe mental ill health (SMI) experience profound health inequalities, with increased rates of long-term physical health problems and behavioral risk factors such as smoking (1). There are concerns that the COVID-19 pandemic will disproportionately impact on the health of people with SMI, increasing already existing health inequalities (2). Around 40% of people with SMI smoke (3), compared to the current smoking prevalence in England of around 14% (4). High rates of smoking remain an important modifiable risk factor, and there is good evidence that effective interventions can encourage successful quitting (5).

The COVID pandemic has impacted on population health risk behaviours and there has been an increase in quit attempts in the wider UK population, though the impact in population smoking rates remains unclear (6). For example, in England it has been reported that 300,000 people quit smoking (7) during the first months of the pandemic. A national campaign urging people to QuitForCOVID was rolled out but it is uncertain whether people with SMI benefitted from this health promotion campaign. UK population surveys (e.g. (6)) have not been able to study this, and it remains possible that disparities in smoking and quitting have been further entrenched among people with SMI during the pandemic.

Studies describing health risk behaviours among people with SMI provide an opportunity to explore changes during the COVID pandemic. Here we describe change in smoking behaviour and quitting among people with SMI.

## Method

The Closing the Gap (CtG) study is a large (n=10,176) clinical cohort recruited between April 2016 and March 2020. Participants have documented diagnoses of schizophrenia or delusional/psychotic illness (ICD 10 F20.X & F22.X or DSM equivalent) or bipolar disorder (ICD F31.X or DSM-equivalent). The composition of the CtG cohort has previously been described (8), and the data at inception included descriptions of self-reported smoking behavior and e-cigarette use.

We were funded to explore the impact of the COVID pandemic in a sub-section of this clinical cohort and we identified participants for Optimising Well-being in Self-Isolation study (OWLS) (https://sites.google.com/york.ac.uk/owls-study/home). To ensure that the OWLS sub-cohort was representative we created a sampling framework based on gender, age, ethnicity and whether they were recruited via primary or secondary care. OWLS participants were recruited from 17 mental health trusts (in six CRN areas across urban and rural settings in England). People who met the eligibility criteria were contacted by telephone or letter and invited to take part in OWLS.

We explored smoking behaviour and e-cigarette use during COVID. For those reporting smoking or using e-cigarettes, we asked whether this was more, less or about the same since the pandemic restrictions began. Those who smoked also completed the smoking heaviness scale (HSI), derived from the Fagerstrom Test for Nicotine Dependence (9) (i.e. how many cigarettes per day they smoked and how long after waking they had their first cigarette), with categories of low, medium and high.

Changes in smoking status since completing the original CtG survey were established by participant linkage between self-reported smoking status before and after the pandemic restrictions began. We also compared smoking statistics within our SMI population with recent figures drawn from the UK-wide survey of smoking in the general population (4).

Ethical approval was granted by the North West – Liverpool Central Research Ethics Committee (reference 20/NW/0276). All participants consented to take part in this study.

## Results

Between July and December 2020, 367 people with SMI were recruited to OWLS. The mean age was 50.5 (range 20-86) with 51% male and 77.4% white British.

Smoking related behavior is given in Table 1. 27.0% (95% CI 22.6 to 31.7) of people who took part in OWLS reported that they smoked compared to 14.1 % in the general population (4). Participants, who smoked, reported smoking a mean of 17.5 cigarettes per day (minimum of 2 and maximum of 60) compared to 9.1 in the general population (4). With reference to the HSI, 6.9% of people from the general population report smoking within five minutes of waking compared to 54.5% in OWLS. In terms of the Heaviness of Smoking Index, 50.5% of people were moderately addicted to smoking and 23.2% highly addicted.

**Table 1:** Smoking behaviour, nicotine dependence, and e-cigarette use in people with SMI.

| Characteristic | OWLS study (N=376)<br>n, % |
| --- | --- |
| % of people who smoke | 99, 27.0% (95% CI 22.6% - 31.7%) |
| Mean number of cigarettes per day (SD) | 17.5 (IQR: 10-20) |
| % of people who smoked within five minutes of waking<sup>a</sup> | 54, 54.5% (95% CI 44.7% – 64.1%) |
| Heaviness of smoking<sup>a, b</sup> |  |
| Low | 21, 21.2% (95% CI 14.1% – 30.0%) |
| Medium | 50, 50.5% (95% CI 40.8% – 60.2%) |
| High | 23, 23.2% (95% CI 15.8% – 32.2%) |
| % of people who use an e-cigarette | 47, 12.8% (95% CI 9.7% – 16.5%) |

a. % are of smokers. b. % do not add up to 100% due to non-respondents.

Of the people who reported smoking in the OWLS survey, 54.5% (95% CI 44.7 to 64.1) said that they had been smoking more heavily since the pandemic restrictions began and 12.1% (95% CI 6.8 to 19.6) said that they had been smoking less.

355 participants provided smoking data both in OWLS and in a pre-COVID data collection. Longitudinal linkage with pre-COVID data showed that, by self-report, 9.0% (95%CI 6.4to 12.3) of people with SMI had stopped smoking. However, 5.9% (95%CI 3.8 to 8.7) of people had started smoking.

## Discussion

Smoking remains one of the most important modifiable risk factors for reduced life expectancy among people with SMI (1). To our knowledge this is the first study to examine smoking status and smoking behaviour change during COVID for people with SMI, and other smoking population surveys have not been able to study this. Our study complements recent population surveys of mental health (10), which have either not been able to capture the experiences of people with SMI, or have focused on mental health but not health risk behaviours. Compared to when cohort participants were first surveyed, pre-COVID the smoking rate has dropped which is encouraging and suggests there have been successful quit attempts however we cannot be certain whether this change in smoking behaviour is as a result of COVID and the pandemic restrictions. We also noted that smoking intensity increased for people who currently smoke, with more than half reporting smoking more. This suggests that although the pandemic may have prompted some people to change their smoking behavior for those who continued to smoke aspects of the pandemic restrictions may have led to them smoking more.

As anticipated when compared to the general population (4) people were smoking on average about twice as many cigarettes per day as people without SMI and people with SMI were nearly ten times more likely to report smoking within five minutes of waking. This indicates that people with SMI who smoke are likely to have a higher level of nicotine dependence than those who do not have SMI, though we cannot link this to COVID. A limitation of the study is that data on nicotine dependence were not available pre-COVID. However we propose to study temporal trends in smoking and nicotine dependence beyond the COVID pandemic.

We conclude that important disparities between the wider population and people with SMI remain, and that smoking related inequalities have potentially increased since the beginning of the COVID pandemic. It is therefore important that effective quit services are provided for, and responsive to, the needs of people who use mental health services.

## Data Availability

The data that support the findings of this study are available on request from the corresponding author.

## Acknowledgements

We would like to thank the participants in the OWLS study and NHS mental health staff for their support with this study.

## Author contribution

All authors contributed to the study protocol. SC, PH, PS and LW recruited participants to the study. EP and SG took the lead in drafting the manuscript. All authors were responsible for critical review of the manuscript for important intellectual content and all authors read and reviewed the transcript.

## Data availability

The data that support the findings of this study are available on request from the corresponding author, EP.

## References

1. Dregan A, McNeill A, Gaughran F, Jones PB, Bazley A, Cross S, et al. Potential gains in life expectancy from reducing amenable mortality among people diagnosed with serious mental illness in the United Kingdom. PLOS ONE. 2020;15(3):e0230674.

2. Druss BG. Addressing the COVID-19 Pandemic in Populations With Serious Mental Illness. JAMA Psychiatry. 2020;77(9):891–2.

3. Public Health England. 2020. [cited 2021].

4. NHS Digital. Statistics on Smoking, England - 2019 2019.

5. Peckham E, Brabyn S, Cook L, Tew G, Gilbody S. Smoking cessation in severe mental ill health: what works? an updated systematic review and meta-analysis. BMC Psychiatry. 2017;17(1):252.

6. Jackson SE, Garnett C, Shahab L, Oldham M, Brown J. Association of the COVID-19 lockdown with smoking, drinking and attempts to quit in England: an analysis of 2019-20 data. Addiction. 2020.

7. Action on Smoking and Health. ASH Daily News for 4 May 2020. 2020.

8. Mishu MP, Peckham EJ, Heron PN, Tew GA, Stubbs B, Gilbody S. Factors associated with regular physical activity participation among people with severe mental ill health. Soc Psychiatry Psychiatr Epidemiol. 2019;54(7):887–95.

9. Heatherton TF, Kozlowski LT, Frecker RC, Rickert W, Robinson J. Measuring the heaviness of smoking: using self-reported time to the first cigarette of the day and number of cigarettes smoked per day. Br J Addict. 1989;84(7):791–9.

10. O’Connor RC, Wetherall K, Cleare S, McClelland H, Melson AJ, Niedzwiedz CL, et al. Mental health and well-being during the COVID-19 pandemic: longitudinal analyses of adults in the UK COVID-19 Mental Health & Wellbeing study. Br J Psychiatry. 2020:1–8.

